# Mortality associated with third generation cephalosporin-resistance in *Enterobacteriaceae* infections: a multicentre cohort study in Southern China

**DOI:** 10.1101/2020.09.11.20192575

**Authors:** Jiancong Wang, Mouqing Zhou, Therese Hesketh, Evangelos I. Kritsotakis

**Author notes:** Corresponding author:* Dr. Evangelos I. Kritsotakis, Laboratory of Biostatistics, School of Medicine, University of Crete, Greece.

## Abstract

**Background:** Emerging third generation cephalosporin-resistant *Enterobacteriaceae* (3GCR-EB) pose a global healthcare concern. We assessed excess mortality in patients infected with 3GCR-EB compared to patients infected with third-generation cephalosporin-susceptible *Enterobacteriaceae* (3GCS-EB).

**Methods:** The study cohort comprised all inpatients with a community-onset or healthcare-associated infection caused by *Enterobacteriaceae* in three tertiary-care public hospitals in 2017. Excess in-hospital mortality was assessed using competing risk survival models, adjusting for baseline patient characteristics.

**Results:** Of 2,343 study patients (median age 60 years; 45.2% male), 1,481 (63.2%) had 3GCS-EB and 862 (36.8%) 3GCR-EB infection. 494 (57.0%) 3GCR-EB isolates were co-resistant to fluoroquinolones and 15 (1.7%) co-resistant to carbapenems. In-hospital mortality was similar in the 3GCS-EB and 3GCR-EB groups (2.4% vs. 2.8%; p=0.601). No increase in the hazard of in-hospital mortality was detected for 3GCR-EB infections compared to 3GCS-EB infections (sub-distribution hazard ratio [HR] 0.80; 95%CI, 0.41 - 1.55) in multivariable analysis adjusting for patient age, sex, intensive care admission, origin of infection and site of infection. Analysis of cause-specific hazards showed that 3GCR-EB infections significantly decreased the daily rate of hospital discharge (cause-specific HR=0.84; 95%CI, 0.76 - 0.92) thereby leading to lengthier hospitalizations.

**Conclusion:** Third-generation cephalosporin resistance in *Enterobacteriaceae* infection per se was not associated with increased in-hospital mortality in this study. However, 3GCR-EB infections were seen to place significant healthcare burden by increasing the length of hospitalization compared to 3GCS-EB infections.

## INTRODUCTION

Emerging third-generation cephalosporin-resistant *Enterobacteriaceae* (3GCR-EB) pose a global healthcare concern in both hospital and community settings [1-3], particularly those harbouring extended-spectrum beta-lactamases (ESBL) [4, 5]. Over the past decade, a significant increase in 3GCR-EB has been observed globally [4-6]. In Europe, the proportion of bloodstream infections due to 3GC-resistant *Escherichia coli* increased from 14% in 2014 to 15% in 2017 [4]; whereas in the United States, the proportion of healthcare-associated infections (HAIs) due to cephalosporin-resistant *E. coli* increased considerably from 23% in 2014 to 30% in 2017 [7]. In Mainland China, the consumption of third-generation cephalosporins is significantly higher than in Europe and the US, both among inpatient [8] and outpatient populations [9]. Consequently, the proportion of *E. coli* resistant third-generation cephalosporins has reached a high level, ranging from 59% to 63% between 2007 and 2017 [6].

Globally, mortality attributable to antimicrobial resistance (AMR) is a major concern [1, 10-14]. Cassini et al. estimated the attributable mortality of AMR in Europe at 6.44 deaths per 100,000 population, causing 170 disability-adjusted life-years per 100,000 population [10]. Another European multicenter study suggested that patients with bloodstream infection due to 3GCR *E. coli* were 2.5 times more likely to die within 30 days following infection onset than patients with bloodstream infection due to third-generation cephalosporin-susceptible *E. coli* [11]. In Asia, the burden of AMR and its clinical impact remain largely understudied because of limited financial resources and laboratory capacity [3, 15, 16]. Indian national research has reported the odds of mortality were 2.6 times higher for patients infected by multidrug-resistant (MDR) *E. coli* compared with non-MDR *E. coli* [15]. In China, a meta-analysis estimated that, in general, patients infected with AMR pathogens have greater likelihood of mortality (odds ratio, 2.7; 95% CI, 2.2 - 3.3) compared to those with infections caused by antimicrobial susceptible organisms [17]. The mortality burden of 3GCR-EB infections in Africa remains largely unknown.

The National Health Commission of the People’s Republic China announced a national action plan to combat AMR in 2016 and multi-disciplinary collaborations with European partners have been initiated to address the increasing burden of AMR in China [17, 18]. However, there remains limited information on the attributable mortality of infection due to 3GCR-EB in Mainland China. This study aimed to fill this research gap by examining the excess mortality associated with the resistance profile of *Enterobacteriaceae* infection in hospitalized patients. To achieve this, in-hospital mortality in patients infected with 3GCR-EB was compared to that of patients infected with third-generation cephalosporin-susceptible *Enterobacteriaceae* (3GCS-EB) based on surveillance data from three tertiary-care hospitals in Southern China in 2017.

## METHODS

### Settings

Dongguan is an industrial city located in Guangdong province, Southern China, with a population of 8.2 million. Dongguan city had a GDP per capita of 14,950 USD in 2018, equivalent to a high-income area according to the World Bank. In 2014, the city established the Dongguan Nosocomial Infection Surveillance System to organize yearly point prevalence surveys, prospective surveillance of surgical site infection, and prospective antimicrobial resistance surveillance [3]. Data in this study were collected from three tertiary-care public hospitals, comprising 8% of all public/not-for-profit hospitals in the city. Collectively, the study hospitals had 3,972 beds (16% of the city’s total beds), completed 133,150 admissions and accumulated 1,366,756 patient-days in 2017 (14.7% and 18.6% of the city’s totals, respectively).

### Study design

This retrospective observational cohort study included all patients in the three study hospitals, who were admitted with a community-onset infection (COI) or developed a HAI caused by *Enterobacteriaceae* in 2017. Patients of all ages in general wards and intensive care units were eligible for inclusion, if a susceptibility test for third-generation cephalosporin resistance had been performed.

### Data collection and definitions

Data collected for each patient enrolled in the study included: demographics (i.e. age, sex), department of admission, dates of hospital admission and discharge, infection data (i.e. date of onset, origin and site/type), antibiogram data for major antimicrobials (i.e. ciprofloxacin, gentamicin, amikacin, piperacillin-tazobactam, and imipenem), and patient outcome upon discharge from the hospital (ascertained as all-cause death or discharged alive). The definitions of HAI included temporal (>48 hours after admission), clinical and microbiological criteria [3, 19]; otherwise, infections diagnosed within 48 hours of admission were defined as COI. In accordance with the criteria of the European Antimicrobial Resistance Surveillance Network (EARS-Net) protocol [4], we took into account the first infection with *Enterobacteriaceae* isolated from clinical samples during the entire hospitalization.

The following indicator organisms in the *Enterobacteriaceae* family were included for analysis: *E. coli, Klebsiella pneumoniae, Klebsiella spp., Enterobacter spp., Citrobacter spp*., and other *Enterobacteriaceae* species (i.e. *Morganella, Proteus, Providencia, Serratia*, and *Salmonella* species). Microbiological identification and susceptibility testing were performed using VITEK® 2 (BioMérieux, Marcy l’Etoile, France). The breakpoints for minimal inhibitory concentration (MIC) were based on the US National Clinical and Laboratory Standards Institute guidelines (modified version based on M100-28^th^ edition in 2017) [20]. The indicator antimicrobial for third generation cephalosporin resistance was ceftriaxone, with an MIC<=1 defining susceptibility and an MIC >=2 defining resistance.

### Statistical analysis

To assess excess mortality due to 3GCR-EB infection compared to 3GCS-EB infection, we applied competing risk survival models [21]. Time to death in the hospital and up to 30 days from the onset of infection was the primary outcome. Survival times of patients who remained hospitalized more than 30 days following infection onset were censored at 30 days. Discharge alive from the hospital was treated as a competing event [21]. To describe the direct effect of 3GCR-EB infection on the two competing outcomes of interest (i.e. discharge alive and in-hospital mortality), we used cause-specific hazard ratios (csHR) that we estimated semi-parametrically by means of separate Cox models for each outcome, assuming proportional hazards. In this analysis, a lower csHR for discharge alive shows that there is a lower daily probability of being discharged alive, thereby a higher risk for longer stay in the hospital following infection onset. Additionally, we described the relative excess in the overall risk of in-hospital mortality, while accounting for the competing event of being discharged alive, using sub-distribution hazard ratios (sHR) that we estimated semi-parametrically by means of the Fine-Gray model [22]. In all models, we adjusted for potential prognostic effects by age, sex, ICU admission, origin of infection (COI vs HAI) and type of infection. The latter was categorized by site as urinary tract, lower respiratory tract, bloodstream and "other” to enable estimation of prognostic effects for the most frequent types of infection. In a supplementary sensitivity analysis, we examined long-term effects by extending the analysis time to 90 days from infection onset. None of the study variables had missing data, except that survival time could not be calculated for 2 patients (<0.1%) because the date of discharge was unknown. All statistical analyses were performed using STATA version 13 (STATA Corp., College station, TX, USA).

### Ethics

The Chinese Ethics Committee of Registering Clinical Trials approved the study and waived the requirement for patient informed consent (approval no: ChiECRCT20190134).

## RESULTS

In total, of 2,509 patients with an *Enterobacteriaceae* infection were identified in the three study hospitals during 2017. Of those, 2,343 (93.4%) had complete antibiogram data and comprised the study cohort (Figure 1). A 3GCR-EB infection had occurred in 862 (36.8%) of the patients. Of the latter, 353 isolates (40.1%) were resistant to 3GC only, 494 (57.3%) were co-resistant to 3GC and fluoroquinolones and 15 (1.7%) were co-resistant to 3GC and carbapenems, with resistance rates being higher in HAIs than COIs (Table I).

**Figure 1.**
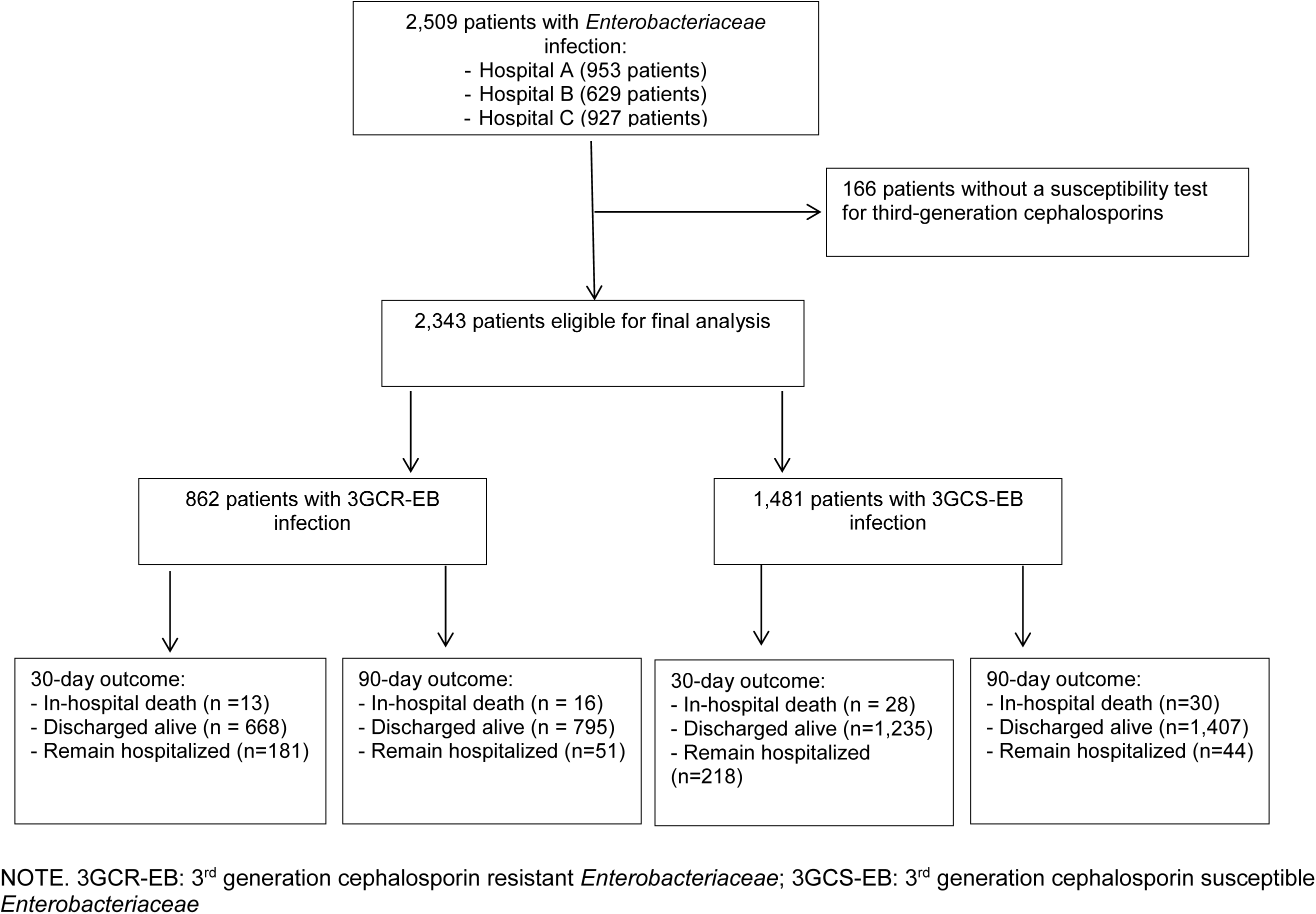
Data extraction flow: Inclusion of patients, patient selection and outcomes

**Table I.** Incidence rates of resistance to 3rd generation cephalosporins and co-resistance to fluoroquinolones and carbapenems in patients with *Enterobacteriaceae* infection

| Antimicrobial resistance group | All patients (N = 2,343) |  | Patients with HAI (N = 449) |  |  | Patients with COI (N = 1,894) |  |  |
| --- | --- | --- | --- | --- | --- | --- | --- | --- |
|  | n | % | n | % | Incidence per 1,000 patient-days (95%CI) | n | % | Incidence per 100 admissions (95%CI) |
| **3GC resistant\*** | 353 | 15.1 | 90 | 20.0 | 0.07 (0.05 – 0.08) | 263 | 13.9 | 0.20 (0.17 – 0.22) |
| **3GC+FQ resistant** | 494 | 21.1 | 113 | 25.2 | 0.08 (0.07 – 0.10) | 381 | 20.1 | 0.29 (0.26 – 0.32) |
| **3GC+CAR resistant** | 6 | 0.3 | 1 | 0.2 | 0.00 (0.00 – 0.00) | 5 | 0.3 | 0.00 (0.00 – 0.01) |
| **3GC+FQ+CAR resistant** | 9 | 0.4 | 5 | 1.1 | 0.00 (0.00 – 0.01) | 4 | 0.2 | 0.00 (0.00 – 0.01) |

3GCR: 3<sup>rd</sup> generation cephalosporin; FQ: fluoroquinolone; CAR: carbapenem; 95%CI: 95% confidence interval.
NOTE. A total of 133,150 admissions with 1,366,756 patient-days occurred during the study period. The table reports incidence proportions for COI (number of resistant COIs per 100 admissions) and incidence density rates for HAI (number of resistant HAIs per 1000 patient-days).
\*3GCR, without known co-resistance to fluoroquinolones or carbapenems.

Table II summarizes baseline characteristics and outcomes of the patients. Median patient age was 60 years (interquartile range 42-74 years, range 0-99 years), 1,058 (45.2%) were males and 115 (4.9%) required adult intensive care at admission. Most patients (80.8%) were admitted with a COI. Urinary tract infection (40.0%), lower respiratory tract infection (20.3%) and bloodstream infection (9.1%) comprised more than two thirds of the infections recorded. Infecting pathogens are summarized by organism and antimicrobial resistance markers in Supplementary Table I. Overall, in-hospital mortality rates at 30 days of follow-up, 90 days of follow-up and hospital discharge were 1.8%, 2.0% and 2.6%, respectively.

**Table II.** Descriptive results of baseline characteristics of the patients and outcomes

| Variables | Overall<br>(n = 2,343) | Enterobacteriaceae infection |  |  |
| --- | --- | --- | --- | --- |
|  |  | 3GCR-EB<br>(n = 862) | 3GCS-EB<br>(n = 1,481) | P* |
| Age, years |  |  |  |  |
| Median (IQR) | 60 (42 - 74) | 60 (42 – 75) | 59 (41 - 74) | 0.985 |
| > 65 years, n (%) | 934 (39.9%) | 343 (39.8%) | 591 (39.9%) | 0.957 |
| Sex |  |  |  |  |
| Male, n (%) | 1,058 (45.2%) | 401 (46.5%) | 657 (44.4%) | 0.311 |
| Department of admission |  |  |  |  |
| Adult intensive care, n (%) | 115 (4.9%) | 42 (4.9%) | 73 (4.9%) | 0.118 |
| Internal medicine, n (%) | 697 (29.8%) | 242 (28.1%) | 455 (30.7%) |  |
| Surgery, n (%) | 1,010 (43.1%) | 399 (46.3%) | 611 (41.3%) |  |
| Pediatrics & Neonatology, (%) | 109 (4.7%) | 43 (5.0%) | 66 (4.5%) |  |
| Other departments, n (%) | 412 (17.6%) | 136 (15.8%) | 276 (18.6%) |  |
| Origin of infection |  |  |  |  |
| Healthcare, n (%) | 449 (19.2%) | 209 (24.3%) | 240 (16.2%) | <0.001 |
| Community, n (%) | 1,894 (80.8%) | 653 (75.8%) | 1,241 (83.8%) |  |
| Site of infection |  |  |  |  |
| Lower respiratory tract, n (%) | 476 (20.3%) | 180 (20.9%) | 296 (20.0%) | 0.106 |
| Urinary tract, n (%) | 937 (40.0%) | 366 (42.5%) | 571 (38.6%) |  |
| Bloodstream, n (%) | 212 (9.1%) | 67 (7.8%) | 145 (9.8%) |  |
| Other, n (%) | 718 (30.6%) | 249 (28.9%) | 469 (31.7%) |  |
| 30-day outcome |  |  |  |  |
| In-hospital death, n (%) | 41 (1.8%) | 13 (1.5%) | 28 (1.9%) | <0.001 |
| Discharged alive, n (%) | 1,903 (81.2%) | 668 (77.5%) | 1,235 (83.4%) |  |
| Remain hospitalized, n (%) | 399 (17.0%) | 181 (21.0%) | 218 (14.7%) |  |
| 90-day outcome |  |  |  |  |
| In-hospital death, n (%) | 46 (2.0%) | 16 (1.9%) | 30 (2.0%) | 0.002 |
| Discharged alive, n (%) | 2,202 (93.9%) | 795 (92.2%) | 1,407 (95.0%) |  |
| Remain hospitalized, n (%) | 95 (4.1%) | 51 (5.9%) | 44 (3.0%) |  |
| Final outcome |  |  |  |  |
| In-hospital death, n (%) | 60 (2.6%) | 24 (2.8%) | 36 (2.4%) | 0.601 |
| Discharged alive, n (%) | 2,283 (97.4%) | 838 (97.2%) | 1,445 (97.6%) |  |
| Length of hospital stay, days |  |  |  |  |
| Patients discharged alive, median (IQR) | 13 (7 – 26) | 15 (8 - 29) | 12 (7 - 24) | <0.001 |
| Patients discharged dead, median (IQR) | 20 (11 - 106) | 30 (15 - 135) | 17 (9 – 61) | 0.056 |

\*Distributions of categorical variables were compared by the Pearson *Chi*-square test. Distributions of continuous variables were compared by the Mann–Whitney U test.

There were 1,481 (63.2%) patients with 3GCS-EB infection and 862 (36.8%) patients with 3GCR-EB infection (Table II). Patients in the 3GCS-EB and 3GCR-EB groups had similar distributions in terms of age, sex, department of admission, and site of infection. However, 3GCR-EB infections were more likely to be healthcare associated (odds ratio = 1.7; p<0.001). Overall, in-hospital mortality was similar in the 3GCS-EB and 3GCR-EB groups (2.4% vs. 2.8%, p=0.601).

In the multivariable survival analysis, there was no statistically significant difference between 3GCR-EB infected patients and 3GCS-EB infected patients in the cause-specific hazards of dying in the hospital within 30 days following infection onset (csHR = 0.74; 95%CI, 0.38 - 1.44; p = 0.379), so the daily hazard of in-hospital death was not increased for 3GCR-EB infected patients (Table III). Similarly, no increase in overall 30-day mortality for patients infected by 3GCR-EB was detected in the analysis of sub-distribution hazards (sHR = 0.80; 95%CI, 0.41 - 1.55; p=0.505). However, 3GCR-EB infection was associated with a statistically significant decrease in the csHR for being discharged alive (csHR = 0.84; 95%CI, 0.76 - 0.92; p<0.001); therefore, the daily hazard of being discharged alive was lower for the 3GCR-EB infected patients, leading to longer hospitalization after being infected by 3GCR-EB, compared to patients with 3GCS-EB infections.

**Table III.** Multivariable competing risk survival analysis for 30-day in-hospital mortality of patients infected by *Enterobacteriaceae*

| Risk factor | Cause-specific hazards |  |  |  |  |  | Sub-distribution hazards |  |  |
| --- | --- | --- | --- | --- | --- | --- | --- | --- | --- |
|  | In hospital death |  |  | Discharge alive |  |  | In hospital death |  |  |
|  | csHR | 95%CI | P | csHR | 95%CI | P | sHR | 95%CI | P |
| **Male sex** | 1.28 | 0.65 - 2.51 | 0.478 | 0.90 | 0.82 - 1.00 | 0.041 | 1.34 | 0.66 - 2.72 | 0.411 |
| **Age > 65 years** | 1.27 | 0.68 - 2.38 | 0.445 | 0.60 | 0.55 - 0.66 | <0.001 | 1.48 | 0.78 - 2.79 | 0.232 |
| **ICU admission** | 2.39 | 1.08 - 5.30 | 0.032 | 0.40 | 0.30 - 0.53 | <0.001 | 3.48 | 1.36 - 8.88 | 0.009 |
| **Origin of infection** |  |  |  |  |  |  |  |  |  |
| Community | 1.00 | - | - | 1.00 | - | - | 1.00 | - | - |
| Healthcare | 1.51 | 0.79 - 2.89 | 0.218 | 0.50 | 0.44 - 0.57 | <0.001 | 1.87 | 0.96 - 3.64 | 0.064 |
| **Site of infection** |  |  |  |  |  |  |  |  |  |
| Lower respiratory tract | 2.73 | 1.02 - 7.29 | 0.045 | 0.83 | 0.72 - 0.95 | 0.009 | 2.92 | 0.98 - 8.72 | 0.055 |
| Urinary tract | 0.41 | 0.11 - 1.49 | 0.176 | 0.85 | 0.76 - 0.96 | 0.006 | 0.45 | 0.12 - 1.71 | 0.244 |
| Bloodstream | 5.27 | 1.93 - 14.42 | 0.001 | 0.90 | 0.76 - 1.06 | 0.207 | 5.40 | 1.96 - 14.87 | 0.001 |
| Other | 1.00 | - | - | 1.00 | - | - | 1.00 | - | - |
| **3<sup>rd</sup> generation Cephalosporin** |  |  |  |  |  |  |  |  |  |
| Susceptible | 1.00 | - | - | 1.00 | - | - | 1.00 | - | - |
| Resistant | 0.74 | 0.38 - 1.44 | 0.379 | 0.84 | 0.76 - 0.92 | <0.001 | 0.80 | 0.41 - 1.55 | 0.505 |

Note: csHR: cause-specific hazard ratio; sHR: sub-distribution hazard ratio; 95% CI: 95% confidence interval

Regarding prognostic effects of other covariates, ICU admission, lower-respiratory tract infection, and bloodstream infection were associated with statistically significant increases in the cause-specific and sub-distribution hazards of 30-day in-hospital mortality (Table III). Of these, ICU admission and lower-respiratory tract infection, but not bloodstream infection, were associated with decreased cause-specific hazards of being discharged alive, leading to longer hospitalization. Advanced age, urinary tract infection, and HAI were associated with increased hospital stay, but not in-hospital mortality.

Consistent results were obtained when the analysis time was extended to 90 days following infection onset (Supplementary Table II). Figure 2 depicts the comparison of the cause-specific cumulative hazards for the two competing events (discharge alive and inhospital mortality) between the 3GCS-EB and 3GCR-EB infection groups.

**Figure 2.**
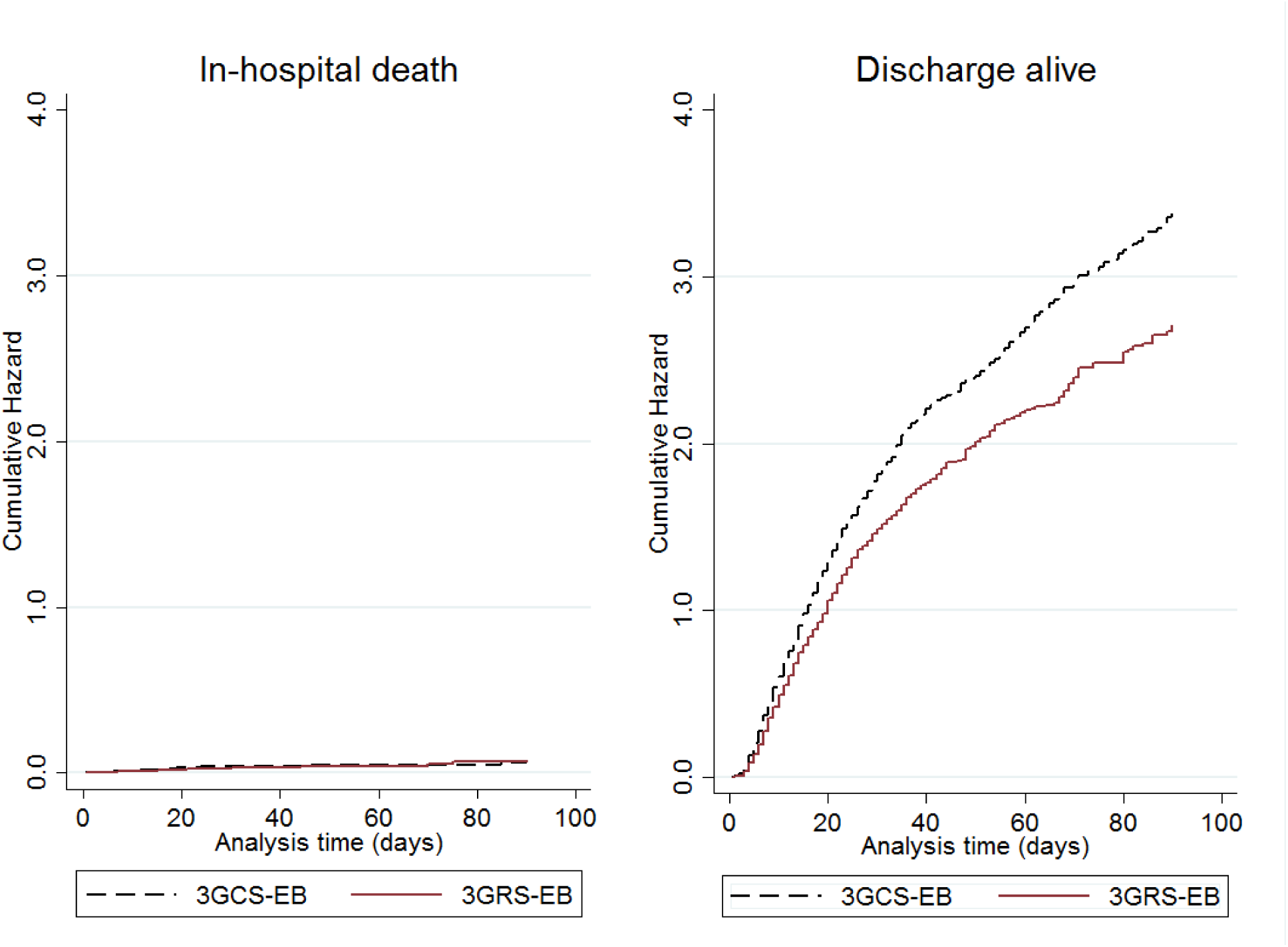
Comparison of cause-specific cumulative hazards of in-hospital death and discharge alive between patients infected by third-generation cephalosporin-susceptible *Enterobacteriaceae* (3GCS-EB) and patients infected by third generation cephalosporin-resistant *Enterobacteriaceae* (3GCR-EB). Cause-specific cumulative hazards were estimated by the Cox model, adjusting for age, sex, intensive care admission, origin of infection and type of infection.Distribution of indicator pathogens resistance to antimicrobial markers in three tertiary-care hospitals (n = 2,777)

## DISCUSSION

In China, a rapid increase of 3GCR-EB infections has been witnessed in the past decade [6]. In this study, we examined the clinical impact of broad-spectrum cephalosporin resistance in hospitalized patients for the first time in Southern China, by comparing mortality hazards between 3GCR-EB and 3GCS-EB infections in a large cohort of patients in three tertiary-care hospitals. Our findings show that the third-generation cephalosporin resistance phenotype in patients infected with *Enterobacteriaceae* was not associated per se with increased risk of in-hospital mortality. This implies that the 3GCR phenotype does not directly add to mortality and/or 3GCR-EB infections can still be managed with appropriate antimicrobial treatment. However, our finding does not diminish the burden of 3GCR-EB infection on patient morbidity and hospital resources [12]. Indeed, our analysis showed that 3GCR-EB infections are associated with decreased daily rate of discharge (alive) from the hospital and thereby led to lengthier hospitalizations compared to 3GCS-EB infections. This is important because prolonged hospitalization increases healthcare costs, increases the risk of other healthcare-associated infections and patient complications, and may increase the risk of transmission of 3GCR-EB to other vulnerable patients.

Existing research on the clinical impact of third-generation cephalosporin resistance in *Enterobacteriaceae* infections has looked at either high risk settings such as the ICU [23, 24] or targeted specific populations such as bacteraemic patients [11, 13, 25] or cancer patients [14]. Few studies have investigated the mortality associated with 3GCR-EB infections in broader acute-care settings [1, 12, 13]. Early single-centre investigations suggested that broad-spectrum cephalosporin resistance is an independent predictor of increased mortality and prolonged length of stay of patients with bacteraemia [13] or other infections [12] caused by *Enterobacter* species. Other, more recent studies observed that the presence of *Enterobacteriaceae* resistance to third-generation cephalosporins was not associated with increased mortality, but did lead to longer length of stay in the ICU [23] and in the wider hospital setting [1]. Conflicting findings regarding the impact of broad-spectrum cephalosporin resistance may be partly explained by variable case-mix (e.g. underlying disease severity, comorbidities and treatment factors). However, it is notable that previous studies [1, 26] disregarded the fact that in-hospital death and discharge (alive) may act as competing outcomes, which is an important factor to consider when analyzing the survival prospects of hospitalized patients [21]. Using appropriate competing risks methodology, we confirmed the lack of excess mortality associated with 3GCR-EB infections, but did note their impact on prolonging hospital stay in multicentre acute-care settings in China.

Our competing risks analysis also allowed a better understanding of the differential clinical impact of other important factors, such as the site and the origin of the infection. We found that bloodstream infection and, to a lesser degree, lower-respiratory tract infection caused by *Enterobacteriaceae* were independently and significantly associated with increased risk of in-hospital death (regardless of their resistance profile). By contrast, lower-respiratory tract infection and, to a lesser degree bloodstream infection, were independently associated with increased risk of prolonged hospital stay (though the effect was not statistically significant for the latter). Urinary tract infections had no effect on hospital mortality, but were associated with significantly increased chances of longer hospitalization. Although not explicitly studied, differential effects by infection site were implied in previous studies on the same topic. For example, Oliveira et al [1] found that a primary site of infection other than UTI was independently associated with all-cause hospital mortality in patients who presented with a 3GCR-EB infection upon hospital admission. Similarly, Kang et al [13] noted that presentation with septic shock and an identified primary site of infection were independent risk factors of 30-day mortality in patients with *Enterobacter* bacteraemia.

Another notable finding from our study is the varying impact by the origin of infection. Although we found no difference in patient mortality between COI and HAI, the latter was significantly and independently associated with decreased probability of being discharged alive (cause-specific HR=0.50; 95%CI, 0.44 - 0.57). This emphasizes the important burden posed by HAIs in prolonging hospitalization. Regarding patient-related risk factors, we found that advanced age (>65 years) was an independent predictor of prolonged hospital stay, but not in-hospital mortality. ICU admission was significantly and independently associated with increased risk of both in-hospital death and prolonged hospitalization.

The main strengths of the present study include its multicentre design with a large sample size and the use of multivariable competing risks models to assess the risk of inhospital mortality. Moreover, the two main exposure groups under comparison (3GCR-EB and 3GCS-EB infected patients) were well balanced in terms of the distribution of important confounders, including age, sex, ICU admission, and site of infection. In addition, there were very few multidrug resistant isolates in our study (only 9 isolates were co-resistant to fluoroquinolones and carbapenems); thus, our analysis is unlikely to have been complicated by complex multi-resistance patterns and pertains specifically to the third-generation cephalosporin resistant phenotype. However, there are potential limitations that we should acknowledge. Data on time-varying confounders such as underlying disease severity and antibiotic therapy were not available in this study and we cannot exclude entirely the possibility that residual confounding may still be present. Moreover, we only looked at inhospital mortality and did not follow up the patients after discharge from the hospital. This might potentially result in informative censoring biasing the results of the Cox models, but it is difficult to assess the magnitude or direction of such bias if it exists.

In conclusion, this investigation of the clinical impact of 3GCR-EB infections in broad acute-care settings in Southern China found no excess risk of in-hospital mortality associated with the 3GCR phenotype in patients with COI or HAI caused by *Enterobacteriaceae*. Nevertheless, 3GCR-EB infections were associated with an increased risk of prolonged hospitalization, thereby placing an important burden on patient morbidity and hospital care.Acknowledgements

The authors thank Dongguan Nosocomial Infection Control and Quality Improvement Centre, and departments of infection control of the three tertiary-care hospitals for providing data for this study.

## Data Availability

The data that support the findings of this study are not publicly available due to their containing information that could compromise the privacy of research participants and study sites. The data are available from the authors, upon reasonable request.

## Author contributions

Conception and design: JW, MZ, and EIK. Acquisition of data: MZ. Data management: JW. Statistical modelling: EIK. Analysis and interpretation of data: JW, MZ, TH and EIK. Drafting the manuscript: JW and EIK. Critical revision for important intellectual content and approval for submission: JW, MZ, TH and EIK. All authors had full access to the study dataset and take responsibility for the integrity of the data presented.

## Transparency declarations

The authors declare that they have no conflict of interest relevant to this study.

---

**Supplementary Table I.**
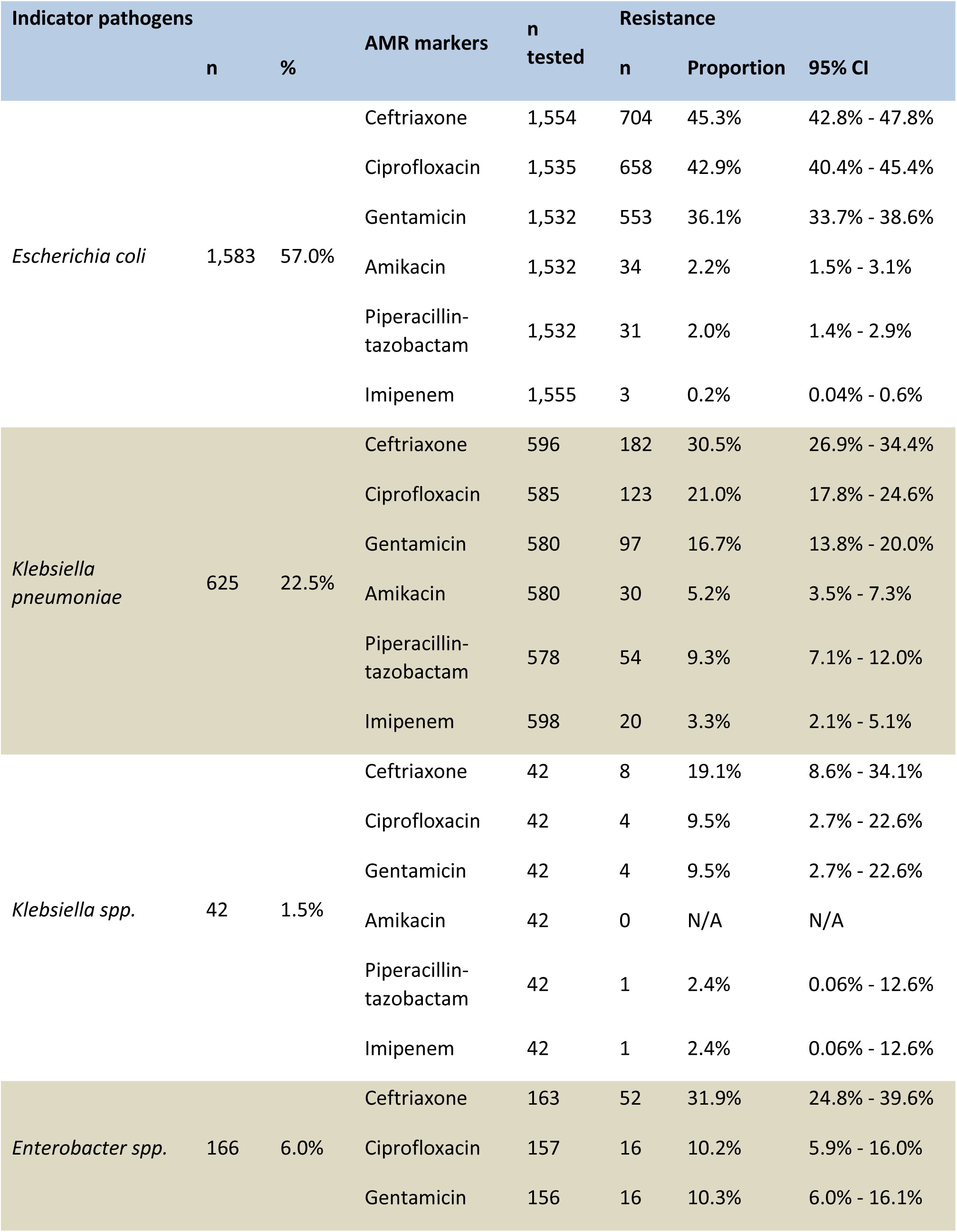
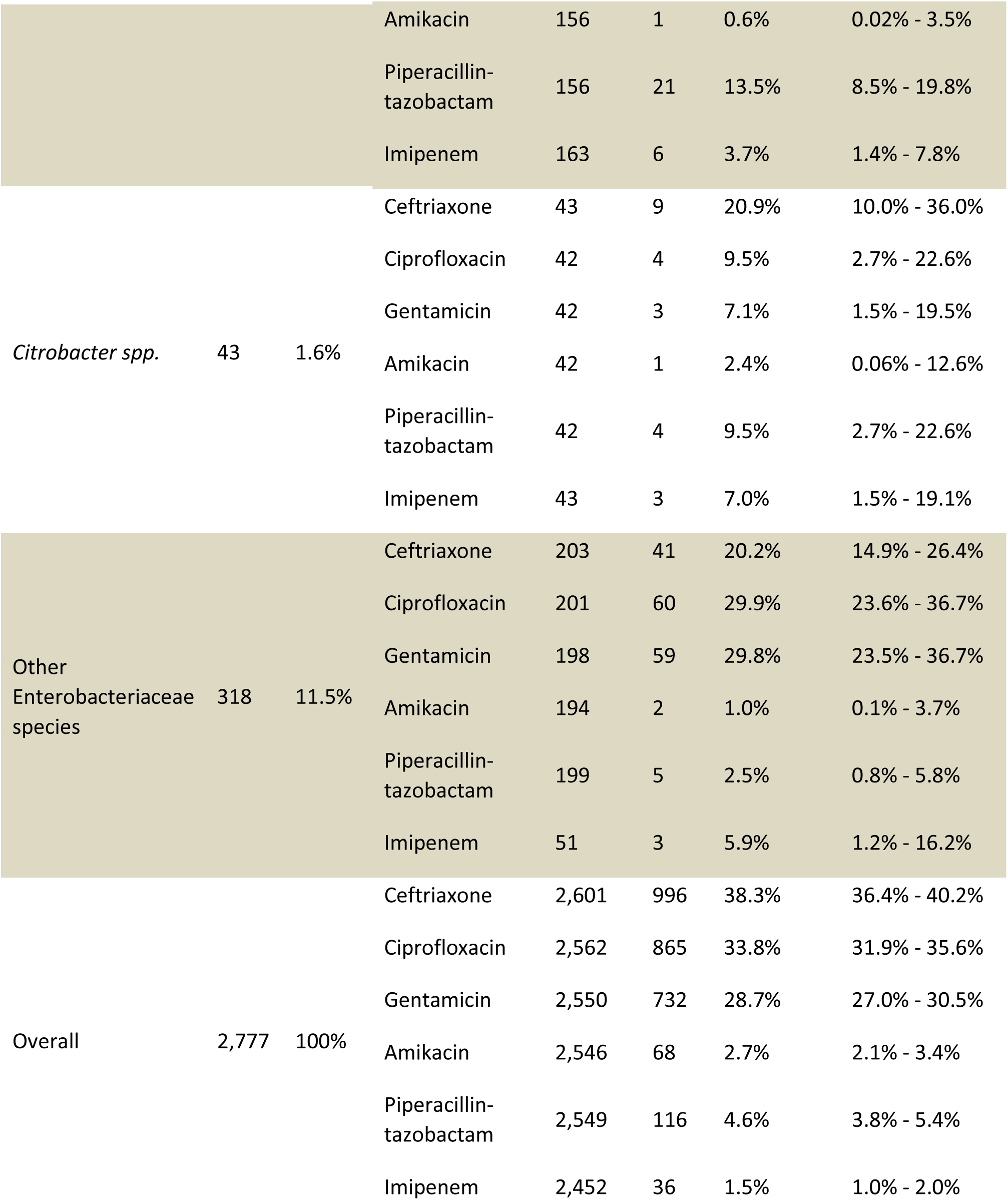
Distribution of indicator pathogens resistance to antimicrobial markers in three tertiary-care hospitals (n = 2,777)

**Supplementary Table II.** Multivariable competing risk survival analysis for **90-day** in-hospital mortality of patients infected by *Enterobacteriaceae*

| Risk factor | Cause-specific hazards |  |  |  |  |  | Sub-distribution hazards |  |  |
| --- | --- | --- | --- | --- | --- | --- | --- | --- | --- |
|  | In hospital death |  |  | Discharge alive |  |  | In hospital death |  |  |
|  | csHR | 95%CI | P | csHR | 95%CI | P | sHR | 95%CI | P |
| **Male sex** | 1.40 | 0.74 - 2.68 | 0.303 | 0.91 | 0.83 - 0.99 | 0.036 | 1.49 | 0.75 - 2.97 | 0.254 |
| **Age > 65 years** | 1.36 | 0.75 - 2.45 | 0.311 | 0.64 | 0.58 - 0.69 | <0.001 | 1.56 | 0.86 - 2.85 | 0.145 |
| **ICU admission** | 1.92 | 0.88 - 4.18 | 0.102 | 0.44 | 0.35 - 0.55 | <0.001 | 2.98 | 1.21 - 7.34 | 0.018 |
| **Origin of infection** |  |  |  |  |  |  |  |  |  |
| Community | 1.00 | - | - | 1.00 | - | - | 1.00 | - | - |
| Healthcare | 1.57 | 0.85 - 2.89 | 0.150 | 0.53 | 0.47 - 0.59 | <0.001 | 2.09 | 1.13 - 3.87 | 0.019 |
| **Site of infection** |  |  |  |  |  |  |  |  |  |
| Lower respiratory tract | 2.48 | 0.99 - 6.20 | 0.051 | 0.80 | 0.70 - 0.91 | 0.001 | 2.79 | 1.02 - 7.60 | 0.045 |
| Urinary tract | 0.44 | 0.14 - 1.43 | 0.173 | 0.85 | 0.77 - 0.95 | 0.004 | 0.49 | 0.15 - 1.62 | 0.241 |
| Bloodstream | 5.09 | 1.98 - 13.05 | 0.001 | 0.89 | 0.76 - 1.05 | 0.162 | 5.06 | 1.97 - 12.99 | 0.001 |
| Other | 1.00 | - | - | 1.00 | - | - | 1.00 | - | - |
| **3<sup>rd</sup> generation Cephalosporin** |  |  |  |  |  |  |  |  |  |
| Susceptible | 1.00 | - | - | 1.00 | - | - | 1.00 | - | - |
| Resistant | 0.80 | 0.43 - 1.48 | 0.483 | 0.84 | 0.77 - 0.92 | <0.001 | 0.90 | 0.49 - 1.67 | 0.748 |

Note: csHR: cause-specific hazard ratio; sHR: sub-distribution hazard ratio; 95% CI: 95% confidence interval

